# Blood acceleration metric: A potential surrogate for fractional flow reserve to assess the functional severity of carotid artery stenosis

**DOI:** 10.64898/2026.11.16.26367829

**Authors:** Maria Margherita Lovera, Vasileios Charitatos, Diane de Zélicourt, Linus Dubs, Stefano Buoso, Carl Moritz Zipser, Susanne Wegener, Sebastian Winklhofer, Hatem Alkadhi, Vartan Kurtcuoglu

## Abstract

Purely morphological assessment of an arterial stenosis is not sufficient to adequately evaluate its functional significance. The metric of fractional flow reserve (FFR) is the gold standard for assessing coronary artery stenoses. However, the risks associated with catheterization and induction of hyperemia needed to measure FFR impede its application to cerebral vessels. Approaches based on a combination of computed tomography (CT) with computer simulations have been proposed to address this issue, yielding an estimate of fractional flow reserved (FFR_CT_). While promising, they are time-consuming and difficult to integrate into established clinical workflows. Here, we propose a new, easily derivable parameter termed the blood acceleration metric (BAM). It incorporates readily measurable morphological and hemodynamic features, such as minimum stenosis diameter and peak systolic velocity, to assess the functional severity of carotid artery stenoses. Preliminary assessment on 27 mild to moderate stenoses indicates a strong negative correlation (R=-0.85) between BAM and FFR_CT_. BAM should be further evaluated as a potential surrogate for FFR and, thereby, as a possible predictor of stroke risk.

## Introduction

Fractional flow reserve (FFR) is the gold standard for evaluating the functional severity of a coronary artery stenosis.^1^ In practice, FFR is estimated as the ratio of the intravascular pressure distal of the stenosis to the proximal one under hyperemic conditions, requiring catheterization for the pressure measurements and administration of a vasodilator such as adenosine to induce hyperemia.^2^ However, these interventions are associated with non-negligible risks (4% risk of transient ischemic attack or stroke, 1% risk of permanent neurological deficit)^3^ which impede the application of FFR to cerebral arteries. To mitigate these risks, the estimation of fractional flow reserve based on computed tomographic angiography (CTA) and computer simulation of blood flow has been proposed as an alternative (FFR_CT_).^4-6^ Arteries of interest are reconstructed from the volumetric CTA data and the associated blood flow and pressure distributions are modeled using computer simulations, allowing for the calculation of FFR at any desired location. Several studies have shown good agreement between computations and catheter-based measurements in the coronaries and in the cerebral and carotid arteries.^4, 6^ While FFR_CT_ shows promise as means to triage patients and minimize unnecessary catheterization, it also requires intricate computational simulations of blood flow that are time consuming and computer resource hungry, making it difficult to integrate into established clinical workflows.

Here, we propose the blood acceleration metric (BAM) as an easily calculated, clinically well-integrable surrogate for FFR_CT_. BAM combines hemodynamic data (e.g., based on duplex ultrasound, DUS) and morphological information (such as from CTA) to assess the functional severity of carotid artery stenosis. To evaluate the correspondence of BAM with FFR_CT_, we employed computational fluid dynamics (CFD) to retrospectively estimate FFR_CT_ in 27 mild to moderate internal carotid artery (ICA) stenoses. A strong correlation between BAM and FFR_CT_ was demonstrated over a large range of stenosis degrees. Given the benefit of FFR_CT_ in the assessment of coronary stenosis severity^6^, BAM appears as a promising functional biomarker for the evaluation of ICA severity.

## Materials and Methods

### Patient selection

We retrospectively screened the radiologic database of the University Hospital Zurich for CTA protocols of patients admitted between 08.06.2019 and 03.11.2019 with suspected stroke. Patients showing at least mild unilateral ICA stenosis according to NASCET criteria^7^ (as described in the CTA reports) were included in the study, regardless of clinical outcome. The study was approved by the ethics committee of the canton of Zurich (BASEC-Nr 2019-02428). Patients gave general consent for research upon hospital admission.

We identified 60 carotid bifurcations with complete DUS reports. Of these, we excluded 25 due to insufficient image quality or imaging artifacts (n=9), dissection (n=2), occlusion (n=1), history of endarterectomy (n=4), history of stenting (n=1), or stenosis-free ICA (n=8). ICAs with stenoses over 70% (n=8) were also excluded, since there already is clinical consensus on the management of symptomatic patients with severe stenoses^7^, and since there the accuracy of DUS peak systolic velocity (PSV) measurement is significantly lower than in moderately stenosed arteries. In total, we included 27 carotid bifurcations with ICA stenosis from 19 different patients. 18 were classified as having ‘mild’ ICA stenosis (10-49% diameter reduction) and 9 as having ‘moderate’ ICA stenosis (50-69% diameter reduction), based on the DUS reports according to NASCET criteria.

### Extraction of carotid geometry and calculation of FPR

We extracted three-dimensional (3D) carotid geometries from the 27 CTA image sets. Using DUS PSV and CFD, we calculated blood flow velocity and pressure distribution at peak systole in these vessels and computed FFR_CT_. The corresponding workflow is illustrated in Figure 1 and detailed in the Supplement. We then compared the performance of three metrics of stenosis severity against FFR_CT_, namely two purely anatomical measures and BAM, the newly proposed metric (Table 1). We note that output of our CFD model has been validated against DUS measurements in previous work^8^.

**Table 1:** Parameters used to quantify stenosis severity.

| Parameter | Definition | Terms |  |
|---|---|---|---|
| % diameter reduction<sup>7</sup> | $\left(1-\frac{D_{sten}}{D_h}\right)\cdot 100\%$ | *D<sub>sten</sub>* | minimum stenosis diameter |
|  |  | *D<sub>h</sub>* | diameter of the closest healthy vessel segment distal to the stenosis |
| % area reduction | $\left(1-\frac{A_{sten}}{A_h}\right)\cdot 100\%$ | $A_{sten}$ | minimum cross-sectional area in the stenosis |
|  |  | $A_h$ | cross-sectional area of the closest healthy vessel segment distal to the stenosis |
| BAM | $\frac{\nu U_{PSV_{CFD},ICA}}{D_{sten}^{2}}$ | *ν* | blood kinematic viscosity (here 3.8 cSt) |
|  |  | $U_{PSV_{CFD},ICA}$ | PSV in the ICA as calculated by CFD |
| FFR<sub>CT</sub> | $\frac{P_d}{P_p}$ | *P<sub>d</sub>* | distal pressure |
|  |  | *P<sub>p</sub>* | proximal pressure |

**Figure 1:**
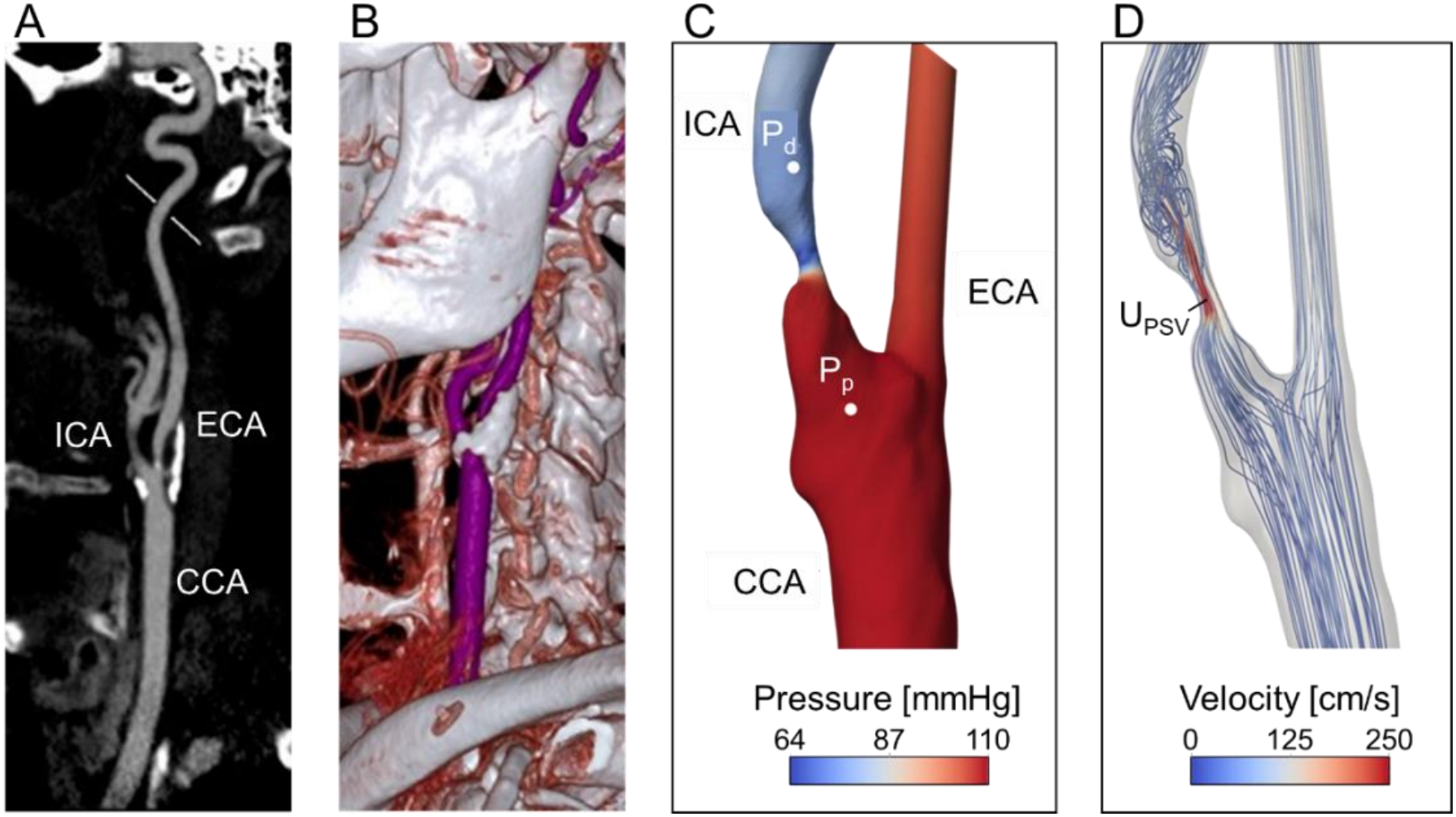
Illustration of the main steps of the study workflow. A: CTA image showing the common, internal and external carotid arteries (CCA, ICA and ECA, respectively), B. 3D rendering of the CTA images. C & D: reconstructed carotid bifurcation geometry and computed velocity and pressure distribution at peak systole. Pp: proximal pressure. Pd: distal pressure. UPSV: peak systolic velocity.

## Results

Figure 1C-D exemplifies flow and pressure distributions within a carotid bifurcation. Blood flow in the stenosis-free ECA is rather uniform and features only minimal pressure drop along its path. In the ICA, pressure drops sharply in the stenosis and recovers only partially distal to it. At the same time, blood is accelerated and reaches its highest velocity in the stenosis.

FFR_CT_ values for the 27 stenoses included in the study and the corresponding parameters of stenosis significance to be evaluated are shown in Figure 2. Data points are color-coded based on stenosis severity according to the NASCET criteria. As illustrated in Figure 2A, the metric of diameter reduction correlates only moderately with FFR_CT_, our reference for functional severity of the stenoses (Pearson correlation coefficient R=-0.60). For example, the three stenoses with the lowest FFR_CT_ do not all show consistently high diameter reduction values; they range between 26% and 65%. Also, some of the stenoses with moderate diameter reduction (black symbols) have FFR_CT_ values comparable to or higher than those of some of the mild stenoses (white symbols). The metric of area reduction shows better correlation with FFR_CT_ (R=-0.71) but falls short of BAM (R=-0.85).

**Figure 2:**
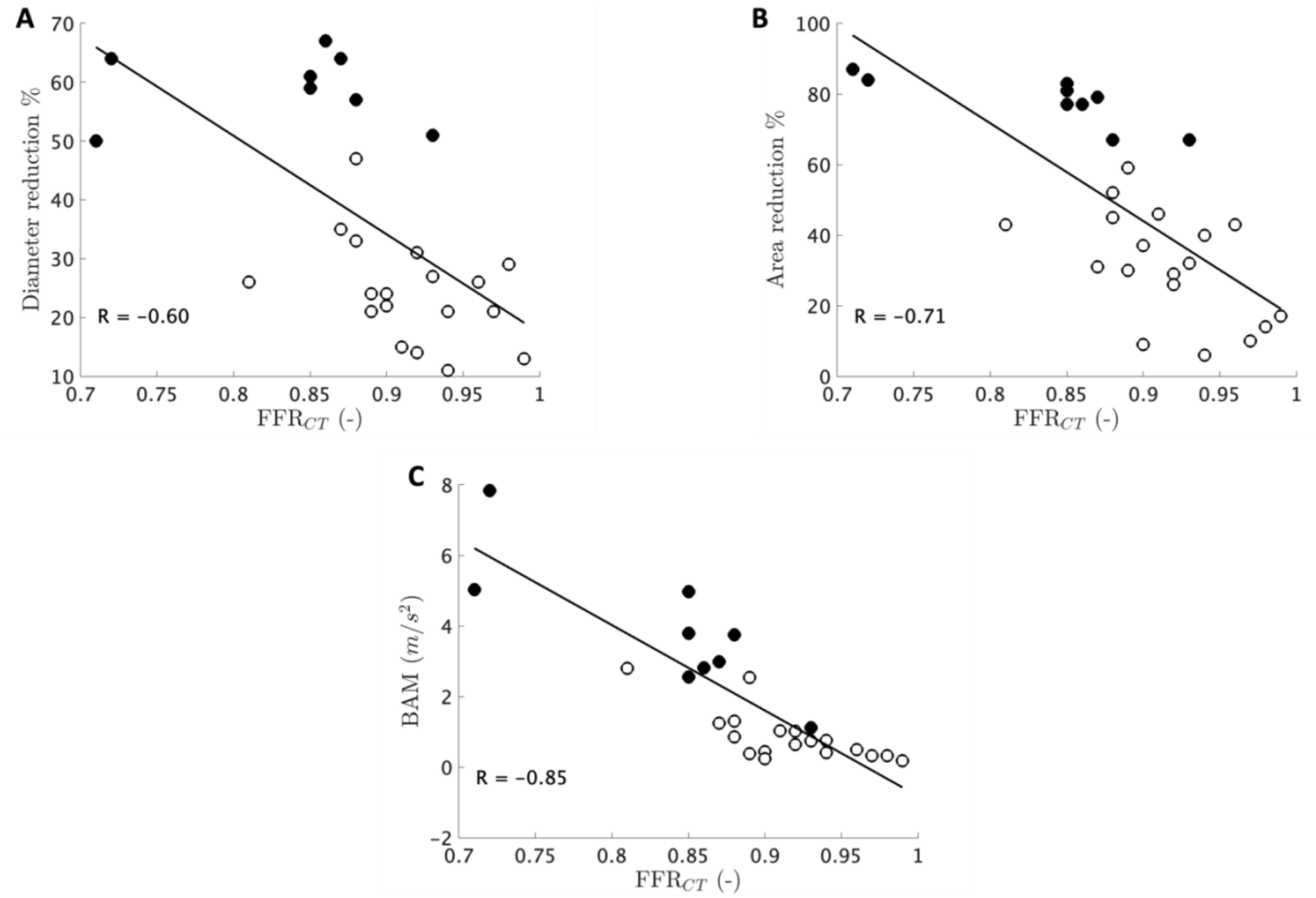
Correlation of FFR with A: Percentage diameter reduction, B. Percentage cross-sectional area reduction, C: Blood acceleration metric (BAM). All correlations were statistically significant with p-values for the two-sided t-test < 0.001. White and black symbols denote, respectively, mild (<50%) and moderate (50-69%) stenoses according to the NASCET criteria.

## Discussion

Our results reiterate the known limited correlation between the functional severity and purely anatomical measures of stenosis.^9^ Miao et al. reported R=-0.53 for the correlation between degree of stenosis and fractional flow measured by catheterization.^10^ Comparably low values were reported by Liu et al. between minimal stenotic area and FFR_CT_ (R=0.55).^5^ Another proposed metric of stenosis severity, the carotid artery blood flow rate, showed better correlation (R=0.68)^4^ with FFR_CT_, but did not reach the level of BAM.

While both stenosis degree and blood flow rate are associated with the functional severity of a stenosis, neither of them provides the complete picture. According to the Hagen-Poiseuille law, the pressure drop along a cylindrical pipe (such as an artery) is (i) proportional to the velocity and viscosity of the fluid flowing through it and (ii) inversely proportional to the pipe’s cross-sectional area. Proceeding by analogy, we defined BAM as the product of blood viscosity and PSV divided by the square of the minimum stenosis diameter (Table 1). Our results indicate a much stronger correlation between FFR_CT_ and BAM (R=-0.85) than with either diameter or area reduction (figures 2A and 2B).

BAM can be determined within seconds with a calculator based on noninvasively acquired clinical data. Measurement of both minimal stenosis diameter and PSV are often part of the clinical workflow for the evaluation of carotid artery stenoses. We used one common value of blood viscosity for all evaluated bifurcations. For patients on blood thinners or those known to have conditions affecting blood viscosity, specific values should be used.

In conclusion, our preliminary results indicate that BAM may be a promising potential surrogate for FFR_CT_ in evaluating the functional severity of carotid artery stenoses. Our findings justify the need for prospective studies to explore the effectiveness of BAM in predicting stroke risk.

## Data Availability

All data produced in the present study are available upon reasonable request to the authors.

