## Supplementary Methods for "Blood acceleration metric: A potential surrogate for fractional flow reserve to assess the functional severity of carotid artery stenosis"

#### Anatomical reconstruction

Three-dimensional (3D) anatomical models of the carotid bifurcations were segmented and reconstructed from the computed tomographic angiography (CTA) images using Syngo.via (Siemens Healthineers, Munich, Germany). We included the common carotid artery (CCA) starting 2-3 cm above the subclavian artery, the internal carotid artery (ICA) up to the carotid canal, and an approximately 5 cm long segment of the external carotid artery (ECA) excluding any side branches. Further flow extensions were added to the inlets and outlets to allow the flow to develop by the time it reached the arterial geometry. This approach has been validated by Hoi et al. under the condition that the length of the extension is at least three times larger than the CCA diameter.[1]

#### Computational fluid dynamics simulations

We generated computational grids for each anatomy using the open-source software GMSH[2] (version 4.2.1) and a vessel diameter-adaptive approach to ensure adequate resolution of the narrowest vessel segments. Grid sizes ranged from 800 000 to 2 000 000 tetrahedral elements, with element sizes between 0.08 mm and 0.5 mm. Grid independence was verified in a preliminary set of simulations by successive refinement of the computational grid until ICA peak systolic velocity (PSV) varied by <10% between consecutive grids.

We conducted blood flow simulations using the open-source FEniCS finite element computing software[3-5] (version 2019.1.0). The unsteady three-dimensional incompressible Navier-Stokes equations were solved as described previously.[6] Blood was modeled as an incompressible Newtonian fluid with density $\rho = 1060\ \frac{kg}{m^3}$ and dynamic viscosity of $\mu = 0.004\ Pa \cdot s$ . The simulations were conducted under subject-specific peak systolic flow conditions. The blood velocity profile at the inlet cross-section of the CCA was assumed to follow a parabolic profile (Poiseuille flow) with a peak velocity equal to the clinically measured PSV:

$$v_{CCA}(x, y) = v_{PSV,CCA} * \left(1 - \frac{x^2 + y^2}{r_{CCA}^2}\right) \quad (1)$$

where $x$ and $y$ are the Cartesian coordinates of a point on the CCA inlet cross-section, $v_{CCA}(x, y)$ is the blood velocity at that location perpendicular to the CCA cross-section, $v_{PSV,CCA}$ is the CCA PSV measured by doppler ultra-sound (DUS), and $r_{CCA}$ is the CCA radius.

A no-slip boundary condition was applied along the vessel walls. Since we only considered one time point, namely peak systole, arterial wall motion due to flow pulsations was neglected. Finally, two-element Windkessel models[7] were coupled to the distal ends of ICA and ECA segments to account for the downstream microcirculation resistance (peripheral resistance) and capacitance:

$$Q = \frac{P}{R_i} + C_i \frac{dP}{dt} \qquad i = ICA, ECA \quad (2)$$

where $Q$ is flow rate, $R_i$ and $C_i$ are the downstream microcirculatory resistance (peripheral resistance) and capacitance of either the ICA or the ICA, respectively, and $dP/dt$ is the change in blood pressure over time. The ICA and ECA downstream resistances, $R_{ICA}$ and $R_{ECA}$, respectively, are calculated based on the patient-specific mean systemic resistance $R_1$. We define $R_1$ as $R_1 = \Delta p / Q_{mean}$, where $\Delta p = 80\ mmHg$ and $Q_{mean} = \pi * r_{CCA}^2 * V_{PSV,CCA}$ is the patient-specific mean flowrate. $R_{ICA}$ and $R_{ECA}$ are then determined by allometric scaling based on their relative radius as:

$$R_{ICA} = R_1 * \frac{\left[\left(\frac{r_{ECA}}{r_{ICA}}\right)^2 + 1\right]}{\left(\frac{r_{ECA}}{r_{ICA}}\right)^2} \quad (3)$$

$$R_{ECA} = \frac{R_{ICA}}{\left(\frac{r_{ECA}}{r_{ICA}}\right)^2} \quad (4)$$

where $r_{ECA}$ and $r_{ICA}$ are the ECA and ICA radii, respectively. Since we considered only time point, the arterial capacitances $C_{ICA}$ and $C_{ECA}$ are considered constant for our simulations and are inversely proportional to the resistances $R_{ICA}$ and $R_{ECA}$.

**Extraction of anatomical parameters**

In the postprocessing phase, the smallest cross-sectional area of the stenosis, denoted as $A_{sten}$, is derived automatically. This is accomplished by computing the surface integrals for the different cross-sectional areas found along the stenosis and selecting the minimum value from these. Given the minimum cross-sectional area's highly irregular and non-circular geometry, the corresponding stenosis diameter, $D_{sten}$, is then determined by averaging two representative diameters from this minimum cross-sectional area. The cross-sectional area $A_h$ and diameter $D_h$ of the closest healthy vessel segment distal to the stenosis, are calculated at 2cm away from the stenosis. The proximal pressure $P_p$ and distal pressure $P_d$ are measured 2 cm proximally and distally of the stenosis, respectively. The $FFR_{CT}$ is then defined as $P_d / P_p$ .

### Supplementary Results

Supplementary Table S1: Anatomical and functional measures of stenosis severity for the 27 carotid bifurcations with ICA stenosis included in the study.

| Case number | Diameter reduction (%) | Area reduction (%) | $FFR_{CT}$ (-) | BAM ($m/s^2$) |
|---|---|---|---|---|
| **1** | 50 | 87 | 0.71 | 5.03 |
| **2** | 21 | 6 | 0.94 | 0.41 |
| **3** | 21 | 30 | 0.89 | 0.38 |
| **4** | 61 | 83 | 0.85 | 2.55 |
| **5** | 24 | 37 | 0.90 | 0.44 |
| **6** | 13 | 17 | 0.99 | 0.18 |
| **7** | 64 | 79 | 0.87 | 2.99 |
| **8** | 22 | 9 | 0.90 | 0.23 |
| **9** | 33 | 45 | 0.88 | 0.85 |
| **10** | 47 | 52 | 0.88 | 1.30 |
| **11** | 51 | 67 | 0.93 | 1.11 |
| **12** | 21 | 10 | 0.97 | 0.32 |
| **13** | 26 | 43 | 0.96 | 0.50 |
| **14** | 15 | 46 | 0.91 | 1.03 |
| **15** | 64 | 84 | 0.72 | 7.84 |
| **16** | 27 | 32 | 0.93 | 0.73 |
| **17** | 11 | 40 | 0.94 | 0.76 |
| **18** | 57 | 67 | 0.88 | 3.74 |
| **19** | 31 | 29 | 0.92 | 1.01 |
| **20** | 35 | 31 | 0.87 | 1.24 |
| **21** | 59 | 81 | 0.85 | 4.98 |
| **22** | 67 | 77 | 0.86 | 2.82 |
| **23** | 29 | 14 | 0.98 | 0.32 |
| **24** | 24 | 59 | 0.89 | 2.53 |
| **25** | 59 | 77 | 0.85 | 3.79 |
| **26** | 26 | 43 | 0.81 | 2.80 |
| **27** | 14 | 26 | 0.92 | 0.64 |

$FFR_{CT}$: estimated fractional flow reserve based on computed tomographic angiography (CTA) and computer simulation of blood flow; BAM: Blood acceleration metric.
